# Reservoir population dynamics and pathogen epidemiology drive pathogen genetic diversity, spillover, and emergence

**DOI:** 10.1101/2020.08.19.20178145

**Authors:** Christopher H. Remien, Scott L. Nuismer

## Abstract

When several factors align, pathogens that normally infect wildlife can spill over into the human population. If pathogen transmission within the human population is self-sustaining, or rapidly evolves to be self sustaining, novel human pathogens can emerge. Although many factors influence the likelihood of spillover and emergence, the rate of contact between humans and wildlife is critical. Thus, for those pathogens inhabiting wildlife reservoirs with pronounced seasonal fluctuations in population density, it is broadly recognized that spillover risk also varies with season. What remains unknown, however, is the extent to which seasonal fluctuations in reservoir populations influence the evolutionary dynamics of pathogens in ways that affect the likelihood of emergence. Here, we use mathematical models and stochastic simulations to show that seasonal fluctuations in reservoir population densities lead to seasonal increases in genetic variation within pathogen populations and thus influence the waiting time for mutations capable of sustained human-to-human transmission. These seasonal increases in genetic variation also lead to elevated risk of emergence at predictable times of year.

## Introduction

Genetic changes are known to affect the transmissibility of infectious diseases [1, 2]. To cause disease, the pathogen must be able to infect and replicate in humans. Pathogen emergence—sustained human-to-human transmission—further requires that the pathogen be sufficiently transmissible in humans, with basic reproductive number *R*_0_ > 1 [3, 4]. A pathogen with an initial *R*_0_ < 1 in the human population could emerge as a result of adaptive evolution if genetic changes increase *R*_0_ to a value greater than one before short transient chains of transmission stutter to extinction [5–8]. Alternatively, emergence could occur if a pathogen spreads well in human populations because it has adapted to closely related species, or if particular strains carry mutations that happen to spread well within humans by chance alone [9]. In this latter scenario, reservoir populations that harbor genetically diverse pathogen populations are more likely to serve as sources for emergence [10].

Many factors affect pathogen genetic diversity, and thus the likelihood that strains capable of human spillover and emergence circulate within the population [11]. For instance, high mutation rates and short generation times allow pathogenic viruses to rapidly increase genetic diversity, whereas bottlenecks at transmission can result in the stochastic loss of genetic variation [12–16]. All else being equal, we would expect pathogen genetic diversity to be greatest within large reservoir populations that harbor correspondingly large pathogen populations. Interestingly, previous work has demonstrated that animals with fast-paced life history strategies characterized by brief lifespan, early sexual maturity, and a large number of offspring are more likely to serve as zoonotic reservoirs [17]. In many cases, these species are also characterized by large population sizes that provide fertile conditions for high pathogen genetic diversity to be maintained. At the same time, however, these fast-paced species often experience seasonal fluctuations in population size that may cause pathogen genetic diversity to wax and wane on annual or multi-annual timescales. For example, both the multimammate mouse (*Mastomys natalensis;* reservoir host of Lassa virus) and the deer mouse (*Peromyscus maniculatus;* reservoir host of Sin Nombre virus) experience large seasonal fluctuations in abundance linked to rainfall and food availability [18–23]. Because these seasonal shifts in abundance can lead to genetic bottlenecks of not just the reservoir species but also the pathogen, they may play a fundamental role in determining the likelihood and timing of pathogen emergence.

Here, we use mathematical models and stochastic simulations to show how the life history of the reservoir population affects pathogen genetic diversity and the likelihood of spillover and emergence. Factors such as seasonal variation in population abundance that lead to large temporal variations in pathogen prevalence in the reservoir can greatly increase the genetic variation of the pathogen at predictable times of the year. This variation may provide windows of opportunity in which the pathogen is well adapted to human hosts, increasing the likelihood of emergence.

## Methods

To determine how demographic and epidemiological factors influence pathogen genetic diversity and emergence risk, we study two mathematical models. First, using a simple deterministic model we derive expectations for pathogen genetic diversity in a reservoir population with constant size. Next, using a stochastic multistrain model we 1) find an approximation for the waiting time for an emergence-capable strain to become the dominant strain within the population; and 2) numerically simulate the model to determine how seasonality affects strain diversity and emergence risk over time. Together these analyses provide insight into which populations have the highest risk of harboring emergent-capable strains and how this risk varies over time.

### Deterministic model with constant population size

One of the most basic factors affecting pathogen genetic diversity is the pathogen’s population size, which is at least partially determined by the number of reservoir hosts infected by the pathogen. If each infected host harbors on average *ρ* pathogens, then we might expect the number of pathogens to be related to the number of infected hosts by *P* = *ρI_host_* where *P* is the number of pathogens and *I_host_* is the number of infected hosts. In reality, epidemiological dynamics and transmission bottlenecks will affect pathogen diversity [12]; yet because we expect pathogen population size to increase with the number of infected hosts, we can gain qualitative insight into the directionality of changes of parameter values on pathogen diversity by studying a simple model of reservoir host population dynamics. In this vein, we interpret a classical ODE model with constant population size in the context of pathogen genetic diversity. A simple model of reservoir epidemiological dynamics with frequency-dependent transmission is given by:

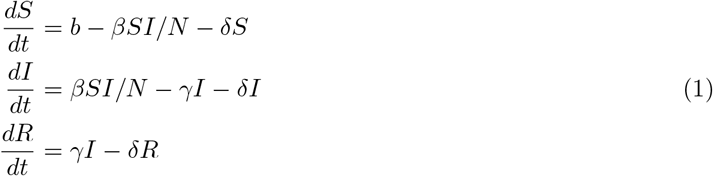

where *S* is the number of susceptible individuals, *I* is the number of infected individuals, *R* is the number of recovered individuals, and *N* = *S* + *I* + *R* is the total population size. Here, *b* is the birth rate, *δ* is the per capita death rate, *β* is the transmission rate, and *γ* is the recovery rate. For this model, the basic reproductive number is *R*_0_ = *β*/(*γ* + *δ*) [24]. At steady state, the number of infected individuals is given by

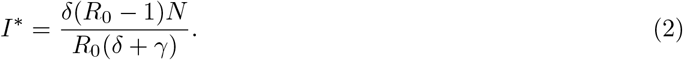

Note that for density-dependent transmission, the steady state value given by equation (2) is identical, but now *R*_0_ = *βN*/(*γ* + *δ*). Because the pathogen population size increases with the number of infected individuals, equation (2) shows how the pathogen population size varies with parameter values. With these basic expectations for how pathogen diversity varies with parameters in hand, we next formulate a stochastic multistrain version of model (1) with a fluctuating birth rate to study how diversity changes over time.

### Stochastic multistrain epidemiological model with fluctuating population size

To understand how fluctuations in reservoir population density impact pathogen diversity and emergence risk for humans, we developed and analyzed a stochastic continuous-time Markov chain multistrain SIR model of the reservoir population. The model is neutral in the sense that individuals infected with each pathogen strain have identical transmission and recovery rates. We assume that there is no coinfection and complete cross-immunity so that after an infected host recovers, it becomes immune to all subsequent reinfections regardless of strain type. Pathogen mutation within infected individuals can generate new strains, and rare strains can stochastically become extinct. In reality, pathogens may exhibit substantial genetic diversity within hosts. Yet if transmission bottlenecks allow only small number of strains to be transmitted from each infected individual, results from our model should hold approximately even in more plausible conditions with multistrain infections. State transitions for the model are given in Table 1 with parameters described in Table 2.

**Table 1:** State transitions and rates at which events occur, given that the current state of the reservoir population is *s* susceptible individuals, *i_j_* individuals infected with strain *j* for *j* = 1,…, *m*, *r* recovered individuals, and 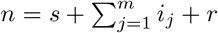 total individuals.

| Event | Transitions | Transition rate |
| --- | --- | --- |
| Birth of susceptible | $s \rightarrow s + 1$ | $b(t)$ |
| Death of a susceptible | $s \rightarrow s - 1$ | $\delta s$ |
| Death of an infected with strain $j$ | $i_j \rightarrow i_j - 1$ | $\delta i_j$ |
| Death of a recovered | $r \rightarrow r - 1$ | $\delta r$ |
| Susceptible infected with strain $j$ | $s \rightarrow s - 1$<br>$i_j \rightarrow i_j + 1$ | $\beta i_j s / n$ |
| Infected with strain $j$ recovers | $i_j \rightarrow i_j - 1$<br>$r \rightarrow r + 1$ | $\gamma i_j$ |
| For $j \neq k$, infected with strain $j$ mutates to strain $k$ | $i_j \rightarrow i_j - 1$<br>$i_k \rightarrow i_k + 1$ | $\mu_{jk} i_j$ |

**Table 2:** Description of parameters in the stochastic mathematical model.

| Parameter | Description |
| --- | --- |
| $\nu$ (individuals) | Average population size |
| $\sigma$ | Clustering of births |
| $\delta$ (day<sup>-1</sup>) | Per capita death rate |
| $\beta$ (day<sup>-1</sup>) | Transmission rate |
| $\gamma$ (day<sup>-1</sup>) | Recovery rate |
| $\mu$ (day<sup>-1</sup>) | Mutation rate for adjacent strains |
| $R_0$ | Basic reproductive number |

The demographic model includes a periodic Gaussian birth rate: *b*(*t*) = *k* exp[−*σ* cos^2^(*πt*/365)]. Other forms of periodicity such as a sinusoidal forcing function or step function could also be used to impose seasonality [25, 26], but Peel et al. [27] found broad support for a periodic Gaussian birth rate in 17 out of 18 datasets on the timing of births in wild animals. The parameter *k* was chosen to ensure an average population size *ν*:

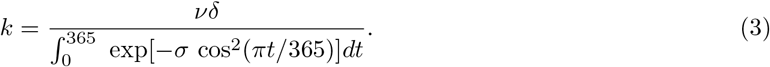

We integrate mutation into our model by assuming a linear strain space with only stepwise mutation at equal rates between all adjacent strains. Specifically, with *n* strains, we assume

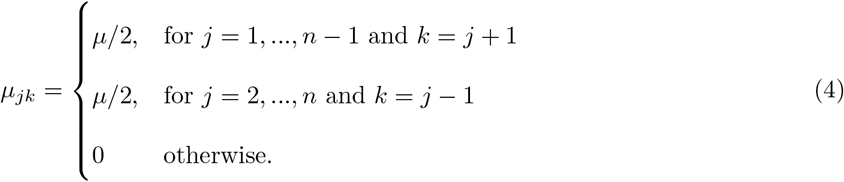

We analyze this basic model in two ways. First, we develop an analytical approximation that calculates the waiting time for an emergence capable mutation to become the dominant (most common) strain within the population. Second, we numerically simulate the model to determine how seasonality affects diversity.

### An approximation for the waiting time to emergence

Here, we wish to quantify the time required for a genetically diverse pathogen to evolve from a state where strains capable of self-sustaining growth within the human population are absent or extremely rare to one where human capable strains dominate. Diffusion time provides such a measure. To calculate the diffusion time, we must first calculate the mean squared displacement, or its square root diffusion length. We found an approximation for diffusion length by considering a random walk in a single infected individual through strain space. Suppose *I*(*x*, *t*) gives the number of individuals infected with strain *x* at time *t*. The average time until a mutation is Δ*t* = 1/*μ*, and because strain space is linear each mutation moves left or right one position in strain space, Δ*x* = 1, with probability 1/2. Taking the limit as Δ*t* and Δ*x* go to zero with 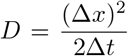 fixed yields the diffusion equation 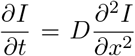 with diffusion coefficient *D* = *μ*/2. The diffusion length is then given by 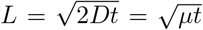. If instead we want to find the time until an average infected individual moves Δ*x* strains away from where it starts, we solve 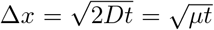 for *t* yielding the diffusion time 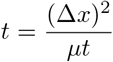.

### Numerical simulations

We simulated the stochastic multistrain model defined in Table 1 in R using the Gillespie algorithm with tau-leaping and a step size of Δ*t* =1 day [28–30]. Specifically, at each time step, the number of times an event with transition rate *f* occurs is Poisson distributed with mean *f*Δ*t*. The transitions are performed, population abundances updated, and the process is repeated. Transitions were bounded to prevent state variables from becoming negative.

To compare with our analytical formula, diffusion length was calculated numerically for each simulation as *L* = (*E*[*I*(*x*, *t*)^2^])^1/2^ where *I*(*x*, *t*) denotes the proportion of infected individuals *x* strains away from the middle strain at time *t*.

## Results

### Which populations are most likely to harbor strains capable of human emergence?

Though the exact formulation varies depending on the diversity metric, pathogen genetic diversity is an increasing function of both the pathogen’s effective population size and its mutation rate. Because pathogen population size is proportional to the infected reservoir population size, increasing the number of infected individuals increases the likelihood of emergence in two ways. First, increasing the number of infected reservoir individuals increases the likelihood of spillover events simply because there are more infected reservoir hosts and thus a higher probability of reservoir-human contact. Second, increasing the number of infected reservoir individuals increases the genetic diversity of the pathogen so that there is a greater chance that an emergent-capable strain is present in the population.

As such, equation (2) highlights how four key parameters influence pathogen genetic diversity and emergence risk. Two of these factors are intrinsic demographic characteristics of the reservoir population (Fig. 1 Panel A). Reservoir populations with larger population sizes and shorter lifespans are predicted to have a higher proportion of infected individuals, and thus higher genetic diversity, all else equal. For a given population size, populations with short lifespans have greater birth rates to maintain constant size, and thus a greater influx of susceptible individuals that can become infected. The other two factors are related to the epidemiology of the pathogen in the reservoir species (Fig. 1 Panel B). Pathogens with larger basic reproductive number and longer duration of infection are predicted to have a higher proportion of infected individuals, and again higher genetic diversity, all else equal. In the extreme case of lifelong infection, a proportion 1-1/*R*_0_ individuals will be infected in this model at steady state. On the other hand, pathogens leading to short durations of infection are predicted to have a low proportion of infected individuals because individuals rapidly recover after becoming infected.

**Figure 1:**
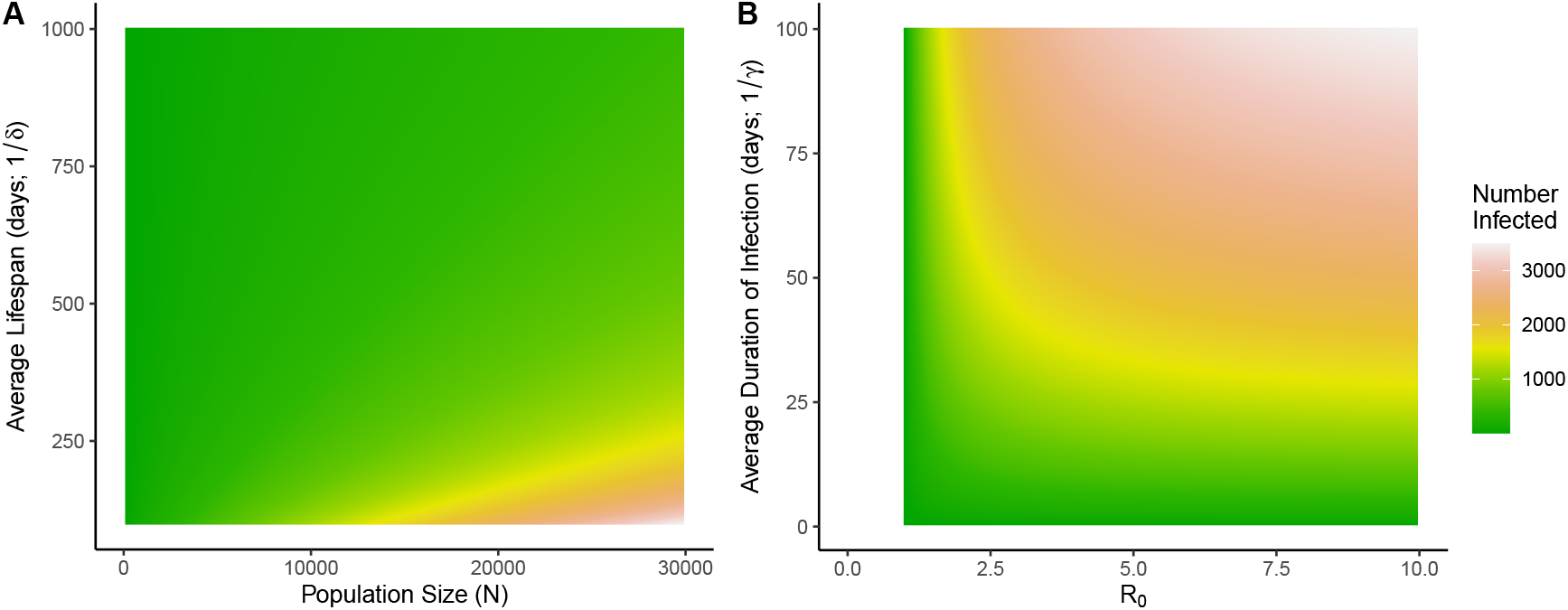
Effect of parameters on the number of infected individuals. Steady-state number of infected individuals in the simple ODE model (equation (2)) for a range of parameter values. Populations with larger numbers of infected individuals are predicted to have greater risk of emergence. A) Reservoir populations with large population sizes (*N*) and short lifespans (1/*γ*) are predicted to have the greatest number of infected individuals. Other parameters are *R*_0_ = 3 and *γ* = 0.05 day^-1^. B) Pathogens with large basic reproductive numbers (*R*_0_) and long durations of infection (1/*γ*) are predicted to have the greatest number of infected individuals. Other parameters are *δ* = 0.01 and *N* = 10, 000.

### How does emergence risk vary over time?

With a baseline expectation for how genetic diversity depends on model parameters, we now explore how emergence risk varies over both multiannual and seasonal time scales.

### Simulating emergence risk in fluctuating populations

We studied seasonal changes in pathogen abundance and genetic diversity by repeatedly simulating our stochastic multistrain epidemiological model (Fig. 2; Table 1). Sample model trajectories over ten years are shown in Fig. 3 Panel A for low/high mutation rates and seasonality. Each realization is unique, but general trends emerge. Seasonal birth pulses increase the abundance of the susceptible population annually (for reference, the reservoir population size is shown in green in Fig. 3 Panel A). This in turn leads to an annual epidemic within the reservoir species with an initially exponentially increasing number of infected individuals. During this initial phase of the epidemic, pathogen diversity rapidly increases. Following this initial phase of the epidemic and subsequent peak, there is a second phase in which the number of infected individuals declines. Each year as the number of infected individuals declines to a critically low level, the pathogen experiences a population bottleneck and the extinction of numerous strains, limiting pathogen genetic diversity. Emergent-capable strains (highlighted in shades of orange in Fig. 3 Panel A) arise stochastically; emergence in the human population is only possible when these strains are present in the reservoir population.

**Figure 2:**
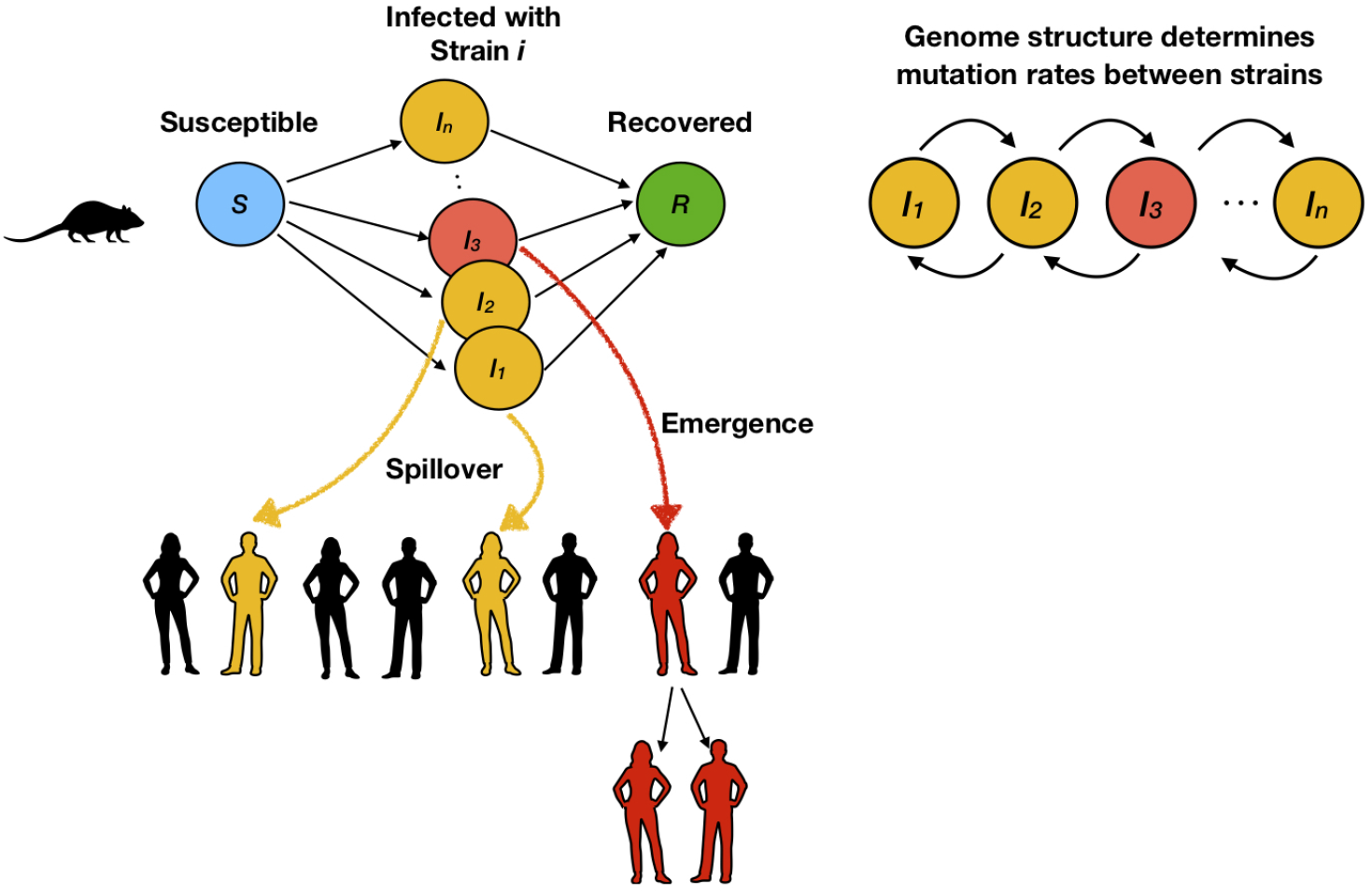
Schematic of the Structure of the Stochastic Model. Our stochastic mathematical model builds on a classical SIR framework. Susceptible reservoir animals can be infected with one of *n* pathogen strains and these animals eventually recover. Pathogen mutation within infected reservoir individuals generates strain diversity within the pathogen population. A subset of pathogen strains are capable of spreading infection to human hosts (spillover), and a subset of these are further capable of sustaining human-to-human transmission (emergence).

**Figure 3:**
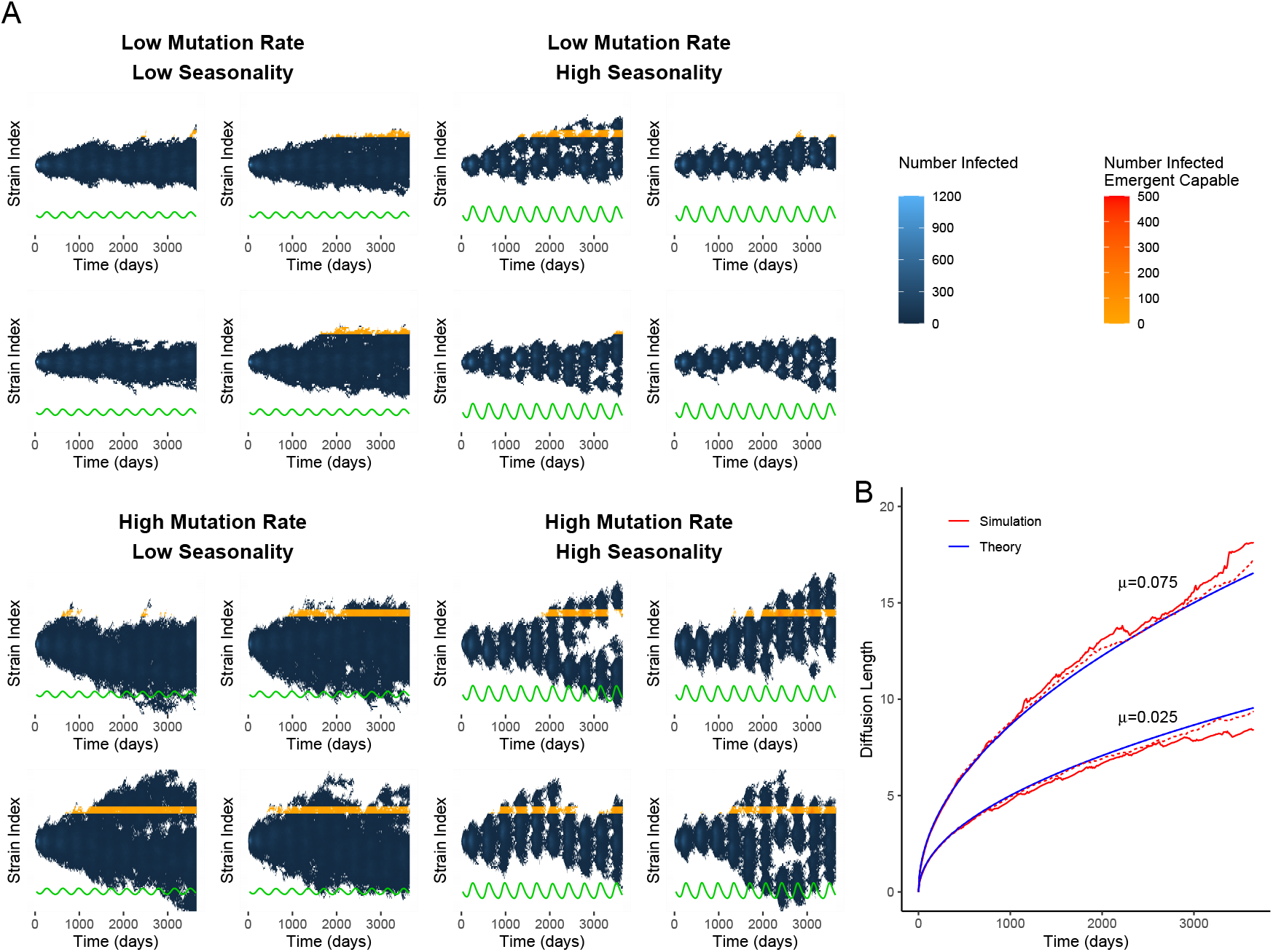
Model Dynamics. A) The number of reservoir individuals infected by each strain plotted over time in shades of blue, for four realizations of the stochastic model for each combination of low/high mutation rate and low/high seasonality. For reference, the reservoir population size is plotted in green. Seasonal increases in population size lead to seasonal epidemics and blooms in pathogen strain diversity. Here, strains from index 70-74 are highlighted as being genetically capable of human emergence in shades of orange. Drift through strain space eventually leads to emergent-capable strains being present in the reservoir population, and their presence and abundance varies seasonally. B) Square root of mean squared displacement—diffusion length—over time. Dashed red lines show the average diffusion length for low seasonality, and solid red lines show the average diffusion length for high seasonality. Averages were calculated from 100 simulations of the model. Over long periods of time, the mutation rate determines the rate at which strain space is explored. Simulations were performed for 100 strains. Initial conditions were set so that there were 25 individuals infected with strain 50, *ν* − 25 susceptible individuals, no individuals infected with other strains, and no recovered individuals. State transitions for the stochastic model are given in Table 1. Parameters are *R*_0_ = 3, *γ* = 0.05 day^−1^, *δ* = 0.01 day^−1^. For low seasonality *σ* =1 while for high seasonality *σ* = 3. The low mutation rate was *μ* = 0.025 day^−1^ and the high mutation rate was *μ* = 0.075 day^−1^. The average population size was *ν* = 10, 000 individuals.

Results from the stochastic simulations confirmed the intuition gained from the deterministic model and show that seasonality in reservoir population abundances is another factor influencing average pathogen diversity (Fig. 4). Such seasonal variation is the result of intrinsic demographic characteristics of the reservoir population (e.g., lifespan, gestation period) as well as the environment in which the particular population is found (e.g., extent of seasonality in rainfall, resource availability, predation). Our stochastic simulations show that populations with large seasonal fluctuations in population size have lower effective population sizes and lower strain diversity on average compared to populations with smaller fluctuations. Sufficiently high seasonality leads to the extinction of the pathogen population.

**Figure 4:**
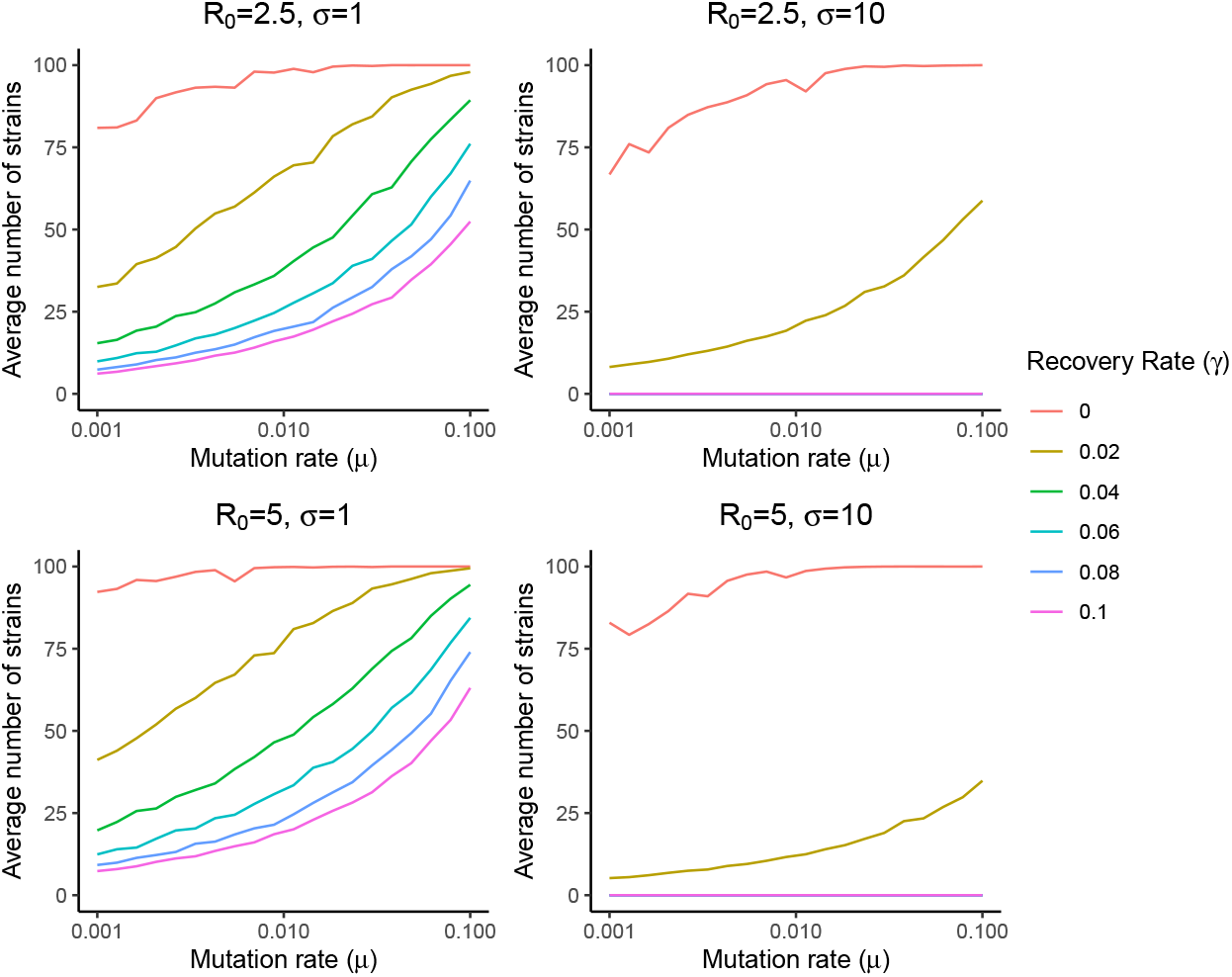
Effect of parameters on average strain diversity. In each panel, the average number of strains present over the course of a year is plotted against mutation rate (*μ*) for a range of recovery rates (y). The top row has *R_0_* = 2.5 and the bottom row has *R*_0_ = 5. The left column has low seasonality (*σ* = 1) and the right column has high seasonality (*σ* = 10). For *σ* = 10, values of *γ* greater than 0.02 day^−1^ led to pathogen extinction for both *R*_0_ = 2.5 and *R*_0_ = 5. Each point is the average of 200 simulations of the stochastic model (Table 1) with 100 strains. For each simulation, twenty five infected individuals were initially seeded randomly to 25 strains. Each simulation was performed for a 25 year burn in period and then sampled for one year. Other parameters were *δ* = 0.01 day^−1^ and *ν* = 10,000 individuals.

### Long-term variation in emergence risk

In the long-term, the time until an emergent strain is present in a particular population depends on the current strain distribution within the population and the mutation rate of the pathogen (Fig. 3 Panel B). With nearest-neighbor mutations and a constant per capita mutation rate, infected individuals follow a random walk across strains. Collectively, this leads to behavior similar to branching diffusion with the diffusion coefficient determined by the mutation rate, *D* = *μ*/2. Our approximation for the diffusion length—a measure of the number of strains the dominant strain of the pathogen is away from the initially dominant strain—is also largely driven by the mutation rate, 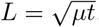. Conversely, the time for an emergent-capable strain to dominate the population can be approximated by 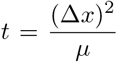 where Δ*x* is the number of strains separating the dominant strain of the population from an emergent-capable strain. Thus a population’s risk horizon—the time until the reservoir population harbors the emergent capable strain—increases with the square of the number of mutations required for the average strain to mutate into an emergent-capable strain. This result shows that over relatively long time-scales, seasonal variation in reservoir abundance plays little role in the likelihood of future emergence. In the next section, however, we will see that over relatively short annual time-scales, reservoir population fluctuations play a central role in the seasonal risk of emergence.

### Seasonal-variation in emergence risk

Emergence depends on the *R*_0_ of the pathogen in the human population. The relationship between pathogen strains and their *R*_0_ values in humans is likely complex and idiosyncratic to the genetics of particular pathogens. Nevertheless certain strains may be capable of sustained human-to-human transmission by chance alone. Because of stochasticity, these emergent-capable strains may not always be present in a given reservoir population but may arise at certain times due to mutation, creating windows of risk in which emergence is possible. Since seasonality increases the variability of strains present annually, it also creates variability in the probability that particular strains that are capable of human-to-human transmission (i.e., those with *R*_0_ > 1 in the human population) are present in the reservoir population. Because our model is neutral (i.e., all strains have the same average transmission and recovery rates and the linear genetic architecture makes all strains equally likely), the likelihood of the emergent-capable strain circulating in the reservoir population is proportional to strain richness—the number of strains present in the population. As such, emergent-capable strains are more likely to be present in the population when strain richness is high.

We quantified how strain richness varies seasonally over a large number of simulations of our stochastic multistrain model (Fig. 5). Here, we were interested in characterizing qualitative changes in the temporal dynamics of strain diversity in response to changes in parameter values. Seasonality leads to annual genetic bottlenecks of the pathogen population which limit strain diversity. These seasonally imposed bottlenecks can create large variations in the number of strains present over the course of a year. We found that the amount of time that peak pathogen prevalence lags peak population abundance depends on the speed of epidemiological dynamics. Pathogens with fast dynamics, characterized by large transmission and recovery rates, cause peak pathogen prevalence to closely track population abundance. In contrast, pathogens with slower dynamics, such as those that cause chronic lifelong infection, cause peak pathogen prevalence to lag behind peak reservoir population abundance. Strain diversity further lags that of pathogen prevalence. Varying the mutation rate, lifespan, and degree of seasonality led to qualitatively similar results, but each of these factors influences the amount of variation in strain diversity over time. High clustering of births in a very brief period of seasonal reproduction leads to larger fluctuations in strain diversity, though as with [27], we found that if population fluctuations are sufficiently large they can lead to extinction of the pathogen.

**Figure 5:**
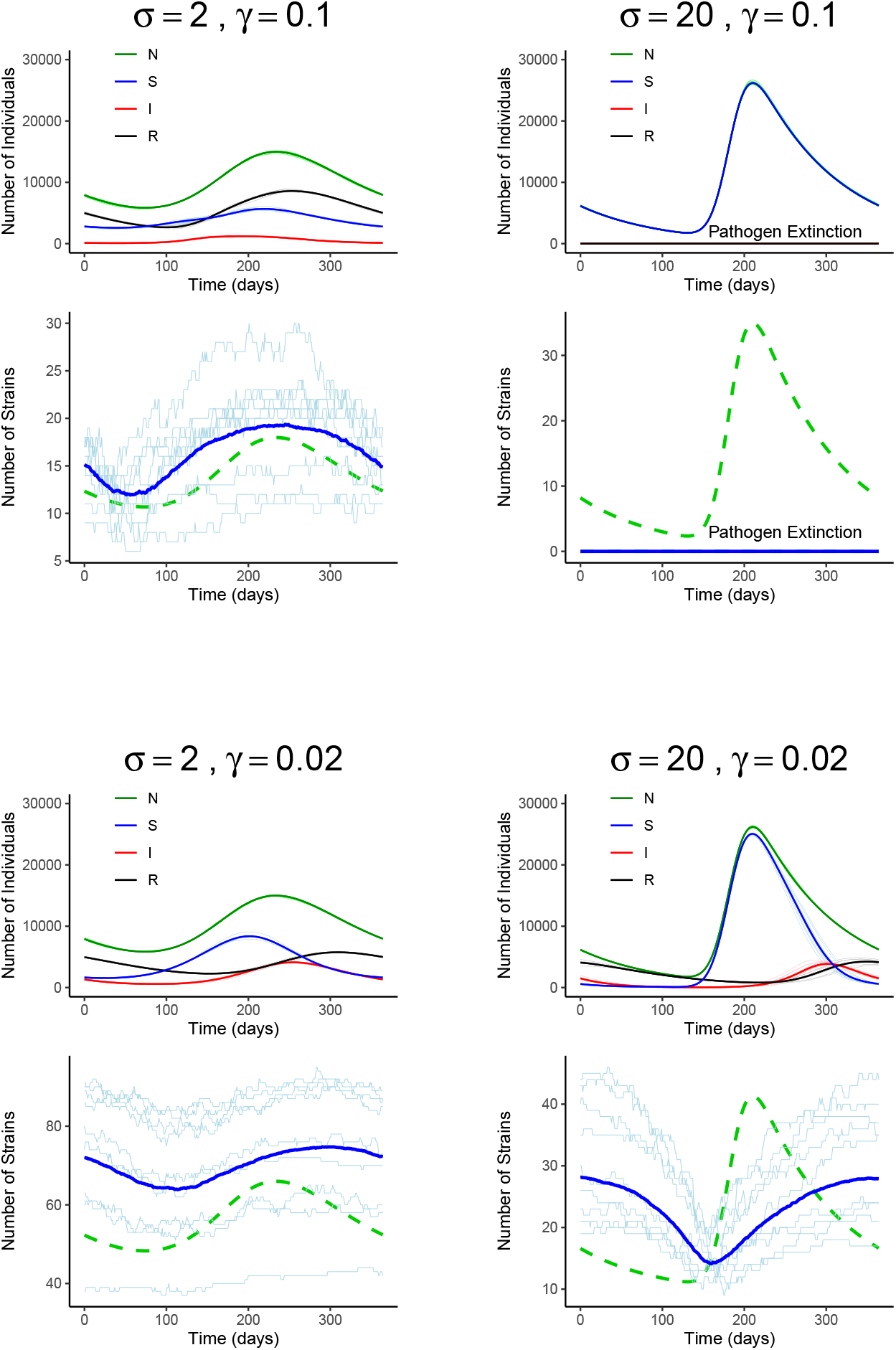
Seasonal dynamics and strain diversity. For each combination of seasonality *σ* and recovery rate *γ*, the top panel shows the average number of susceptible, infected, and recovered individuals over time, and ten sample simulations in light colors. The bottom panels show the average number of strains present (strain richness) over time (dark blue) and ten sample simulations (light blue). For reference, the reservoir population size is shown in green. The reservoir population size and strain richness vary over time due to a time-varying birth rate. Note that the range of the vertical axis varies for the plots of strain richness. Averages are from 100 simulations of the model with 100 strains for each combination of parameter values. For each simulation, twenty five infected individuals were initially seeded randomly to 25 strains. Each simulation was performed for a 25 year burn in period and then sampled for one year. State transitions for the model are given in Table 1. Other parameters were *R*_0_ = 2.5, *μ* = 0.02 day^−1^, *δ* = 0.01 day^−1^, *ν* = 10,000 individuals.

## Discussion

Emergence of a new pathogen in humans requires the culmination of numerous, potentially rare, events [9, 31, 32]. Reservoir animals must not only be infected with the pathogen, but must harbor strains capable of both infecting humans and sustaining human-to-human transmission. Identifying reservoir populations that have such strains circulating within them, or strains similar enough to emergent-capable strains that mutation toward such strains is plausible, will be critically important for identifying reservoir populations with highest potential for human emergence [33–35]. Our results show how demographic characteristics of the reservoir population and the properties of the epidemiology of the pathogen affect pathogen diversity.

Seasonality in reservoir population abundance can greatly influence pathogen diversity over the course of a year, and thus the probability that strains capable of emergence are found in a reservoir population at any point in time. Seasonal epidemics fueled by the births of susceptible individuals can lead to the rapid expansion of strain diversity while bottlenecks induced by population crashes lead to reductions in strain diversity. For pathogens in reservoir species with large seasonal fluctuations in abundance, the risk that emergent-capable strains are circulating lags reservoir population abundance. The duration of lag depends on pathogen dynamics. Our models predict that pathogens with high transmission and recovery rates, such as Lassa virus in *Mastomys natalensis*, have peak risk of emergent-capable strains circulating roughly coincident with peak reservoir population abundance. In contrast, for pathogens with low transmission and recovery rates, such as *Sin Nombre orthohantavirus* in *Peromyscus maniculatus* which is thought to cause lifelong infection in its rodent host, peak risk may lag population abundance by as much as six months. Some support for these predictions has been observed in specific systems. For instance, Adler et al. [23] found a time-lag in prevalence between population peaks of *Peromyscus maniculatus* and peaks in *Sin Nombre orthohantavirus* antibody prevalence and Kallio et al. [36] found that human infections with *Puumala hantavirus* lag the abundance of their rodent host *Myodes glareolus*.

For emergence to occur, reservoir individuals infected with an emergent-capable strain must contact humans (or other animals that can go on to infect humans) in a way that allows transmission. The timing of peak risk of emergence may not be directly tied to peak pathogen abundance and diversity depending on the ecology of the reservoir species and the nature of their interaction with humans. For example, *Mastomys natalensis*, a major reservoir of Lassa virus, primarily interacts with humans during the dry season when it is more likely to be found inside people’s homes, so that peak risk of spillover is in the dry season [19, 37]. Other reservoirs such as bats which can be infected with Hendra virus, Nipah virus, and coronaviruses may have complex life cycles and ecologies which influence risk. For example, environmental conditions that lead to resource scarcity may increase the co-occurance of bats and horses, increasing the risk of transmission of Hendra virus from bats to horses and on to humans [38, 39]. While our models capture the basic dynamics of fluctuating wildlife populations, more detailed models of specific human-wildlife interactions will be required to study how spillover and emergence risk vary seasonally for specific pathogen-reservoir-human population combinations. Pathogen genetics will be an important factor to include in such models.

Our results indicate that pathogen population bottlenecks can lead to the clustering of specific strains within each reservoir host population (i.e., reduced strain variance within each given population). High levels of seasonality tend to lead to populations in which all pathogen strains are closely related. Thus, for species with high levels of variation in abundance, certain reservoir populations may harbor pathogens with high potential for emergence, while the pathogens of other populations may be more benign. In populations with less demographic seasonality, the risk may be more uniform across populations. An implication of this is that demographic seasonality may lead to increased variation of pathogen strains present across space, though the degree to which this manifests depends on migration and other factors specific to the reservoir host ecology. This heterogeneity between populations may be important for identifying the populations of the wildlife reservoir with the greatest potential for harboring emergent-capable pathogen strains.

A more complete understanding of pathogen evolution within their reservoir hosts is required to predict when and where the next pandemic is likely to arise. Here, we have shown that reservoir population dynamics and pathogen epidemiology can affect the genetic variation of pathogens in ways that affect the likelihood of spillover and emergence over time. Models that incorporate the complex interplay between pathogen genetics, reservoir population dynamics, and interactions between the reservoir host and humans will play an important role in forecasting future pandemics and targeting interventions to preempt them.

## Data Availability

NA

## References

[1] Domingo, Esteban. Mechanisms of viral emergence. Vet. Res., 41(6):38, 2010.

[2] Ben Longdon, Michael A Brockhurst, Colin A Russell, John J Welch, and Francis M Jiggins. The evolution and genetics of virus host shifts. PLoS pathogens, 10(11):e1004395–e1004395, 11 2014.

[3] Roy Malcolm Anderson and Robert Mccredie May. The population dynamics of microparasites and their invertebrate hosts. Philosophical Transactions of the Royal Society of London. B, Biological Sciences, 291(1054):451–524, 1981.

[4] Philippe Carmona and Sylvain Gandon. Winter is coming: Pathogen emergence in seasonal environments. PLOS Computational Biology, 16(7):1–16, 07 2020.

[5] Rustom Antia, Roland R Regoes, Jacob C Koella, and Carl T Bergstrom. The role of evolution in the emergence of infectious diseases. Nature, 426(6967):658–661, 12 2003.

[6] N. Arinaminpathy and A. R. McLean. Evolution and emergence of novel human infections. Proceedings of the Royal Society B: Biological Sciences, 276(1675):3937–3943, 2009.

[7] James O. Lloyd-Smith, Dylan George, Kim M. Pepin, Virginia E. Pitzer, Juliet R. C. Pulliam, Andrew P. Dobson, Peter J. Hudson, and Bryan T. Grenfell. Epidemic dynamics at the human-animal interface. Science, 326(5958):1362–1367, 2009.

[8] Edward C. Holmes. The comparative genomics of viral emergence. Proceedings of the National Academy of Sciences, 107(suppl 1):1742–1746, 2010.

[9] Colin R Parrish, Edward C Holmes, David M Morens, Eun-Chung Park, Donald S Burke, Charles H Calisher, Catherine A Laughlin, Linda J Saif, and Peter Daszak. Cross-species virus transmission and the emergence of new epidemic diseases. Microbiology and Molecular Biology Reviews, 72(3):457–470, 09 2008.

[10] John J. Dennehy, Nicholas A. Friedenberg, Robert C. McBride, Robert D. Holt, and Paul E. Turner. Experimental evidence that source genetic variation drives pathogen emergence. Proceedings of the Royal Society B: Biological Sciences, 277(1697):3113–3121, 2010.

[11] Cas Retel, Hanna Märkle, Lutz Becks, and Philine G D Feulner. Ecological and evolutionary processes shaping viral genetic diversity. Viruses, 11(3):220, 03 2019. doi:10.3390/v11030220.

[12] Carl T. Bergstrom, Paul McElhany, and Leslie A. Real. Transmission bottlenecks as determinants of virulence in rapidly evolving pathogens. Proceedings of the National Academy of Sciences, 96(9): 5095–5100, 1999.

[13] Laura A. Shackelton, Colin R. Parrish, Uwe Truyen, and Edward C. Holmes. High rate of viral evolution associated with the emergence of carnivore parvovirus. Proceedings of the National Academy of Sciences, 102(2):379–384, 2005.

[14] Edward C. Holmes. The evolutionary genetics of emerging viruses. Annual Review of Ecology, Evolution, and Systematics, 40(1):353–372, 2009.

[15] David A. Kennedy and Greg Dwyer. Effects of multiple sources of genetic drift on pathogen variation within hosts. PLOS Biology, 16(3):1–17, 03 2018.

[16] Rafael Sanjuán and Pilar Domingo-Calap. Genetic diversity and evolution of viral populations. Reference Module in Life Sciences, pages B978–0–12–809633–8.20958–8, 2019.

[17] Barbara A. Han, John Paul Schmidt, Sarah E. Bowden, and John M. Drake. Rodent reservoirs of future zoonotic diseases. Proceedings of the National Academy of Sciences, 112(22):7039–7044, 2015.

[18] Herwig Leirs, Nils Chr. Stenseth, James D. Nichols, James E. Hines, Ron Verhagen, and Walter Verheyen. Stochastic seasonality and nonlinear density-dependent factors regulate population size in an african rodent. Nature, 389(6647):176–180, 1997.

[19] Elisabeth Fichet-Calvet, Emilie Lecompte, Lamine Koivogui, Barré Soropogui, Amadou Doré, Fodé Kourouma, Oumar Sylla, Stéphane Daffis, Kékoura Koulémou, and Jan Ter Meulen. Fluctuation of abundance and lassa virus prevalence in mastomys natalensis in guinea, west africa. Vector-Borne and Zoonotic Diseases, 7(2):119–128, 2007.

[20] Scott L. Nuismer, Christopher H. Remien, Andrew Basinski, Tanner Varrelman, Nathan Layman, Kyle Rosenke, Brian Bird, Michael Jarvis, Peter Barry, and Elisabeth Fichet-Calvet. Bayesian estimation of lassa virus epidemiological parameters: implications for spillover prevention using wildlife vaccination. bioRxiv, 2019.

[21] M. Andrea Previtali, Erin M. Lehmer, Jessica M. C. Pearce-Duvet, Jeremy D. Jones, Christine A. Clay, Britta A. Wood, Patrick W. Ely, Sean M. Laverty, and M. Denise Dearing. Roles of human disturbance, precipitation, and a pathogen on the survival and reproductive probabilities of deer mice. Ecology, 91 (2):582–592, 2010.

[22] Terry L. Yates, James N. Mills, Cheryl A. Parmenter, Thomas G. Ksiazek, Robert R. Parmenter, John R. Vande Castle, Charles H. Calisher, Stuart T. Nichol, Kenneth D. Abbott, Joni C. Young, Michael L. Morrison, Barry J. Beaty, Jonathan L. Dunnum, Robert J. Baker, Jorge Salazar-Bravo, and Clarence J. Peters. The Ecology and Evolutionary History of an Emergent Disease: Hantavirus Pulmonary Syndrome: Evidence from two El Niño episodes in the American Southwest suggests that El Niño-driven precipitation, the initial catalyst of a trophic cascade that results in a delayed density-dependent rodent response, is sufficient to predict heightened risk for human contraction of hantavirus pulmonary syndrome. BioScience, 52(11):989–98, 11 2002.

[23] Frederick R. Adler, Jessica M. C. Pearce-Duvet, and M. Denise Dearing. How host population dynamics translate into time-lagged prevalence: An investigation of sin nombre virus in deer mice. Bulletin of Mathematical Biology, 70(1):236, 2007.

[24] Matt J Keeling and Pejman Rohani. Modeling Infectious Diseases In Humans and Animals. Princeton University Press, 2008.

[25] R Donnelly, A Best, A White, and M Boots. Seasonality selects for more acutely virulent parasites when virulence is density dependent. Proceedings of the Royal Society B: Biological Sciences, 280 (1751):20122464–20122464, 01 2013.

[26] Courtney L. Schreiner, Scott L. Nuismer, and Andrew J. Basinski. When to vaccinate a fluctuating wildlife population: Is timing everything? Journal of Applied Ecology, 57(2):307–319, 2020.

[27] A. J. Peel, J. R. C. Pulliam, A. D. Luis, R. K. Plowright, T. J. O’Shea, D. T. S. Hayman, J. L. N. Wood, C. T. Webb, and O. Restif. The effect of seasonal birth pulses on pathogen persistence in wild mammal populations. Proceedings of the Royal Society B: Biological Sciences, 281(1786):20132962, 2014.

[28] Daniel T Gillespie. A general method for numerically simulating the stochastic time evolution of coupled chemical reactions. Journal of Computational Physics, 22(4):403–434, 1976.

[29] Daniel T. Gillespie. Exact stochastic simulation of coupled chemical reactions. The Journal of Physical Chemistry, 81(25):2340–2361, 12 1977.

[30] Daniel T. Gillespie. Approximate accelerated stochastic simulation of chemically reacting systems. The Journal of Chemical Physics, 115(4):1716–1733, 2001.

[31] S. Cleaveland, D. T. Haydon, and L. Taylor. Overviews of Pathogen Emergence: Which Pathogens Emerge, When and Why?, pages 85–111. Springer Berlin Heidelberg, Berlin, Heidelberg, 2007.

[32] Raina K Plowright, Colin R Parrish, Hamish McCallum, Peter J Hudson, Albert I Ko, Andrea L Graham, and James O Lloyd-Smith. Pathways to zoonotic spillover. Nature Reviews Microbiology, 15 (8):502–510, 08 2017.

[33] Daniel T Haydon, Sarah Cleaveland, Louise H Taylor, and M Karen Laurenson. Identifying reservoirs of infection: a conceptual and practical challenge. Emerging infectious diseases, 8(12):1468–1473, 12 2002.

[34] Mark Woolhouse, Fiona Scott, Zoe Hudson, Richard Howey, and Margo Chase-Topping. Human viruses: discovery and emergence. Philosophical Transactions of the Royal Society B: Biological Sciences, 367 (1604):2864–2871, 2012.

[35] Kevin M. Bakker, Tonie E. Rocke, Jorge E. Osorio, Rachel C. Abbott, Carlos Tello, Jorge E. Carrera, William Valderrama, Carlos Shiva, Nestor Falcon, and Daniel G. Streicker. Fluorescent biomarkers demonstrate prospects for spreadable vaccines to control disease transmission in wild bats. Nature Ecology & Evolution, 3(12):1697–1704, 2019.

[36] Eva R. Kallio, Michael Begon, Heikki Henttonen, Esa Koskela, Tapio Mappes, Antti Vaheri, and Olli Vapalahti. Cyclic hantavirus epidemics in humans-predicted by rodent host dynamics. Epidemics, 1 (2):101–107, 2009.

[37] Elisabeth Fichet-Calvet, Emilie Lecompte, Lamine Koivogui, Stephane Daffis, and Jan ter Meulen. Reproductive characteristics of mastomys natalensis and lassa virus prevalence in guinea, west africa. Vector Borne and Zoonotic Diseases, 8(1):41–48, 2008.

[38] Raina K Plowright, Hume E Field, Craig Smith, Anja Divljan, Carol Palmer, Gary Tabor, Peter Daszak, and Janet E Foley. Reproduction and nutritional stress are risk factors for hendra virus infection in little red flying foxes (*pteropus scapulatus*). Proceedings of the Royal Society B: Biological Sciences, 275 (1636):861–869, 2008.

[39] Raina K. Plowright, Peggy Eby, Peter J. Hudson, Ina L. Smith, David Westcott, Wayne L. Bryden, Deborah Middleton, Peter A. Reid, Rosemary A. McFarlane, Gerardo Martin, Gary M. Tabor, Lee F. Skerratt, Dale L. Anderson, Gary Crameri, David Quammen, David Jordan, Paul Freeman, Lin-Fa Wang, Jonathan H. Epstein, Glenn A. Marsh, Nina Y. Kung, and Hamish McCallum. Ecological dynamics of emerging bat virus spillover. Proceedings of the Royal Society B: Biological Sciences, 282 (1798):20142124, 2015.

